# Rapid and accurate point-of-care testing for SARS-CoV2 antibodies

**DOI:** 10.1101/2020.11.30.20241208

**Authors:** Sally Esmail, Michael J. Knauer, Husam Abdoh, Benjamin Chin-Yee, Peter Stogios, Almagul Seitova, Ashley Hutchinson, Farhad Yusifov, Tatiana Skarina, Elena Evdokimova, Suzanne Ackloo, Lori Lowes, Courtney Voss, Benjamin D. Hedley, Vipin Bhayana, Ian Chin-Yee, Shawn S-C. Li

**Affiliations:** Departments of Biochemistry, Schulich School of Medicine and Dentistry, Western University, London, Ontario N6G 2V4, Canada; Department of Pathology and Laboratory Medicine, Western University and London Health Sciences Centre, 800 Commissioners Rd E, London, Ontario N6A 5W9, Canada; Divison of Hematology, Western University and London Health Sciences Centre, 800 Commissioners Rd E, London, Ontario N6A 5W9, Canada; Structural Genomics Consortium, University of Toronto, 101 College St, MaRS South Tower, Suite 700, Toronto, Ontario M5G 1L7, Canada; Department of Chemical Engineering and Applied Chemistry, University of Toronto, 200 College St., Toronto, Ontario M5S 3E5, Canada

## Abstract

The COVID-19 pandemic, caused by the severe acute respiratory syndrome coronavirus-2 (SARS-CoV-2), has grown into worst public health crisis since the 1918 influenza pandemic. As COVID-19 continues to spread around the world, there is urgent need for a rapid, yet accurate antibody test to identify infected individuals in populations to inform health decisions. We have developed a rapid, accurate and cost-effective serologic test based on antibody-dependent agglutination of antigen-coated latex particles, which uses ∼5 µl plasma and takes <5 min to complete with no instrument required. The simplicity of this test makes it ideal for point-of-care (POC) use at the community level. When validated using plasma samples that are positive or negative for SARS-CoV-2, the agglutination assay detected antibodies against the receptor-binding domain of the spike (S-RBD) or the nucleocapsid (N) protein of SARS-CoV-2 with 100% specificity and ∼98% sensitivity. Furthermore, we found that the strength of the S-RBD antibody response measured by the agglutination assay correlated with the efficiency of the plasma in blocking RBD binding to the angiotensin converting enzyme 2 (ACE2) in a surrogate neutralization assay, suggesting that the agglutination assay may be used to identify individuals with virus-neutralizing antibodies. Intriguingly, we found that >92% of patients had detectable antibodies on the day of positive viral RNA test, suggesting that seroconversion may occur earlier than previously thought and that the agglutination antibody test may complement RNA testing for POC diagnosis of SARS-CoV-2 infection.

## INTRODUCTION

Development of rapid point-of-care COVID-19 diagnostics for use at the community level remains a top priority in the global response to the COVID-19 pandemic^1^. While capacity for detecting SARS-CoV-2 based on nucleic acid amplification (NAAT) has grown immensely and enabled effective public health responses, serologic testing for virus specific antibodies has not gained the same widespread application, due to concerns over sensitivity, specificity, cost and turn-around time^1,2^. Although NAAT is the current gold standard for diagnosing acute infection, it is not effective in identifying individuals who have recovered from previous infection^3^. Given that approximately 40% of infected individuals remain asymptomatic^1,4,5^, large-scale antibody testing could help better establish the true extent of the COVID-19 pandemic, identifying disease hotspots and high-risk populations to enable more effective isolation and contact tracing^1,3,6^. Moreover, antibody testing may identify individuals with a strong neutralizing antibody response who may be suitable donors for convalescent plasma/serum therapy for the treatment of those with severe symptoms^7^.

To date, a number of antibody tests have been approved for emergency use in the US and Europe. These tests detect the IgG, IgM or IgA antibody against the spike (including the receptor binding domain, RBD) or nucleocapsid (N) protein of the SARS-CoV-2 virus by enzyme-linked immunosorbent assay (ELISA)^3^. ELISA-based antibody tests, which can be qualitative or quantitative, require specialized instruments and are usually performed in a lab by a trained technician. The sensitivity and specificity of different ELISA kits vary widely^8-10^. To enable point-of-care (POC) testing, several rapid diagnostic tests (RDTs) based on lateral flow have been developed. Although the RDTs reduced the time of the antibody test to 10-30 minutes from 2-5 hours (for ELISA), they generally suffer from decreased sensitivity and specificity compared to ELISA-based assays^1,11-13^.

To address the pressing need for a simple, rapid, yet accurate antibody test^1^, we resorted to the tested-and-proven serology method of agglutination that has been used in blood typing and antibody testing^14-16^. We show here that the agglutination of red blood cells (RBCs) or latex particles induced by specific antigen-antibody interaction affords a highly sensitive and accurate assay for SARS-CoV-2 antibodies. We validated the antibody assay based on latex particle agglutination using 169 plasma samples that were tested positive for SARS-CoV-2 by NAAT, 121 samples that were NAAT negative and 100 SARS-CoV-2 naïve plasma samples. The agglutination-based antibody assay produced 100% specificity and 97-98.2% sensitivity. Because this simple assay requires no instrument and generates results in 2 minutes, it has the potential to be used as a POC or at-home test.

## RESULTS

### Agglutination-based serologic testing for SARS-CoV-2 antibodies

Agglutination of RBCs is widely used in blood typing whereas latex particle agglutination assays have been used to detect antibodies against a variety of different viruses. We sought to establish whether either or both approaches could be adapted for SARS-CoV-2 antibody testing. In principle, coating the RBCs or the latex particles with a SARS-CoV-2 specific antigen would enable their respective agglutination by the corresponding antibody (Fig. 1). To explore this notion, we labelled Group O (R2R2) RBCs carrying the D antigen with the spike RBD (S-RBD) or the RNA binding domain of the nucleocapsid (N-RBD) protein through streptavidin-biotin mediated coupling (Figs. S1 & S2, see Materials and Methods for details). Incubating the antigen-coated RBCs with COVID-19^+^ plasma led to robust agglutination whereas the COVID-19^-^ plasma failed to induce RBC agglutination, suggesting that the aggregation of the S-RBD/N-RBD-coated RBCs may be used to detect antibody response to SARS-CoV-2 (Fig. S2).

**Figure 1:**
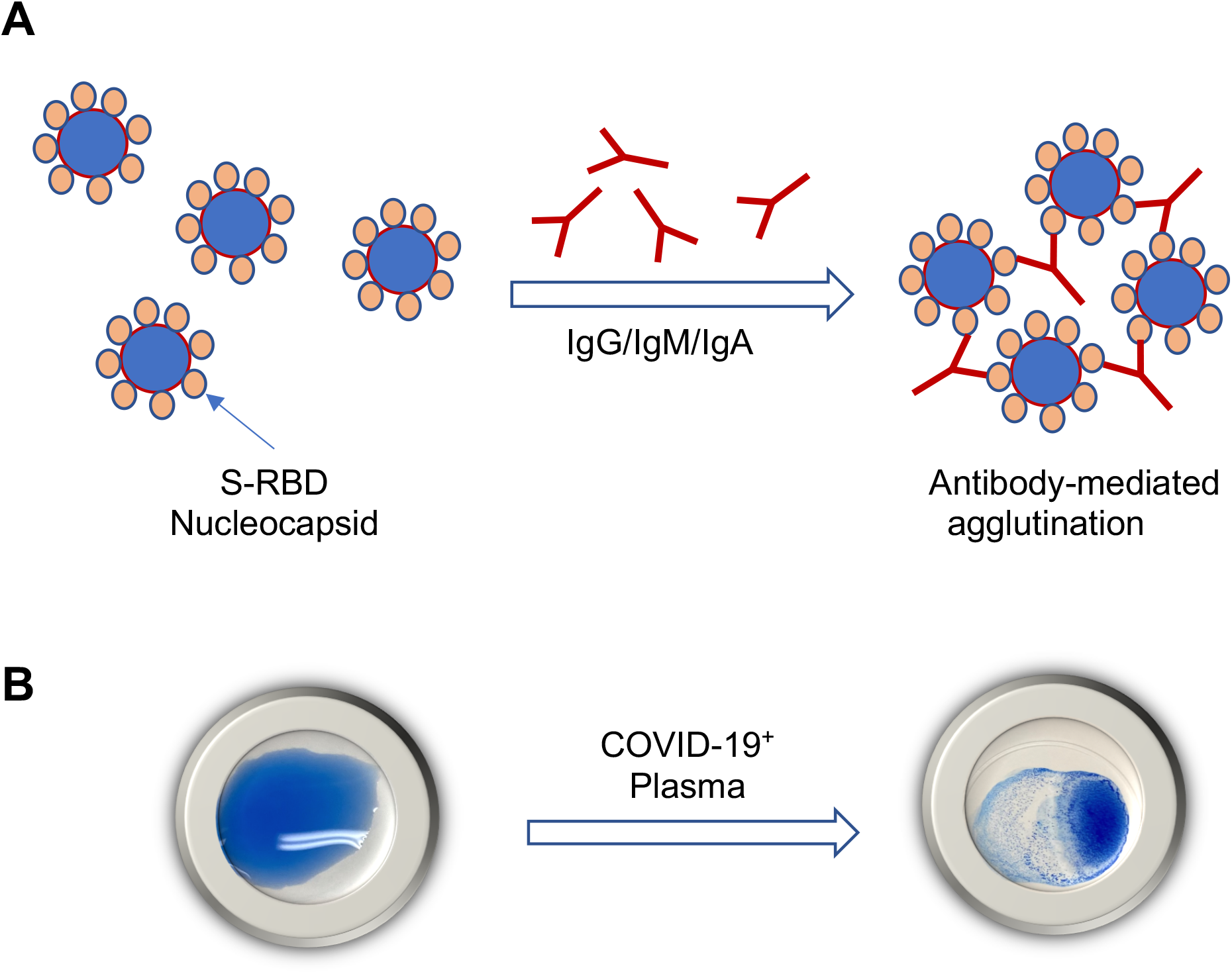
Illustration of the principle of agglutination assay for SARS-CoV-2 antibody testing. (**A**) Latex particles or red blood cells(RBC) are surface-coated with a SARS-CoV-2 antigen, the S-RBD or nucleocapsid (N). Incubation with plasma containing antibodies against the coated antigen would induce agglutination of the latex particles or RBCs. (**B**) A representative image of the agglutination assay using latex beads coated with S-RBD.

To develop a cost-effective agglutination assay, we next investigated if latex particles were a suitable substitute for the RBCs. To this end, we coated latex particles with recombinant S-RBD or the full-length nucleocapsid (N) protein (Fig. S1). The antigen coated latex beads were first tested with a monoclonal anti-S-RBD and a polyclonal anti-nucleocapsid antibody. Upon incubating with the corresponding antibody, the antigen-coated latex particles formed clumps within two minutes. Importantly, the area of clump formation grew larger with increasing antibody concentrations (Fig. S3). Although latex agglutination was commonly used as a qualitative assay, it is possible to determine the degree of agglutination based on the area of clump formation via image analysis. As shown in Fig. 2, the percentage of agglutination for both the S-RBD- and N-coated latex particles increased when an incremental amount of anti-S-RBD or anti-N antibody was added. Fitting the data to Hill’s equation yielded Hill’s coefficient of 1.7 for the former and 1.8 for the latter. This suggests that the antibody-induced agglutination of latex particles is a cooperative event (Fig. 2A&B).

**Figure 2.**
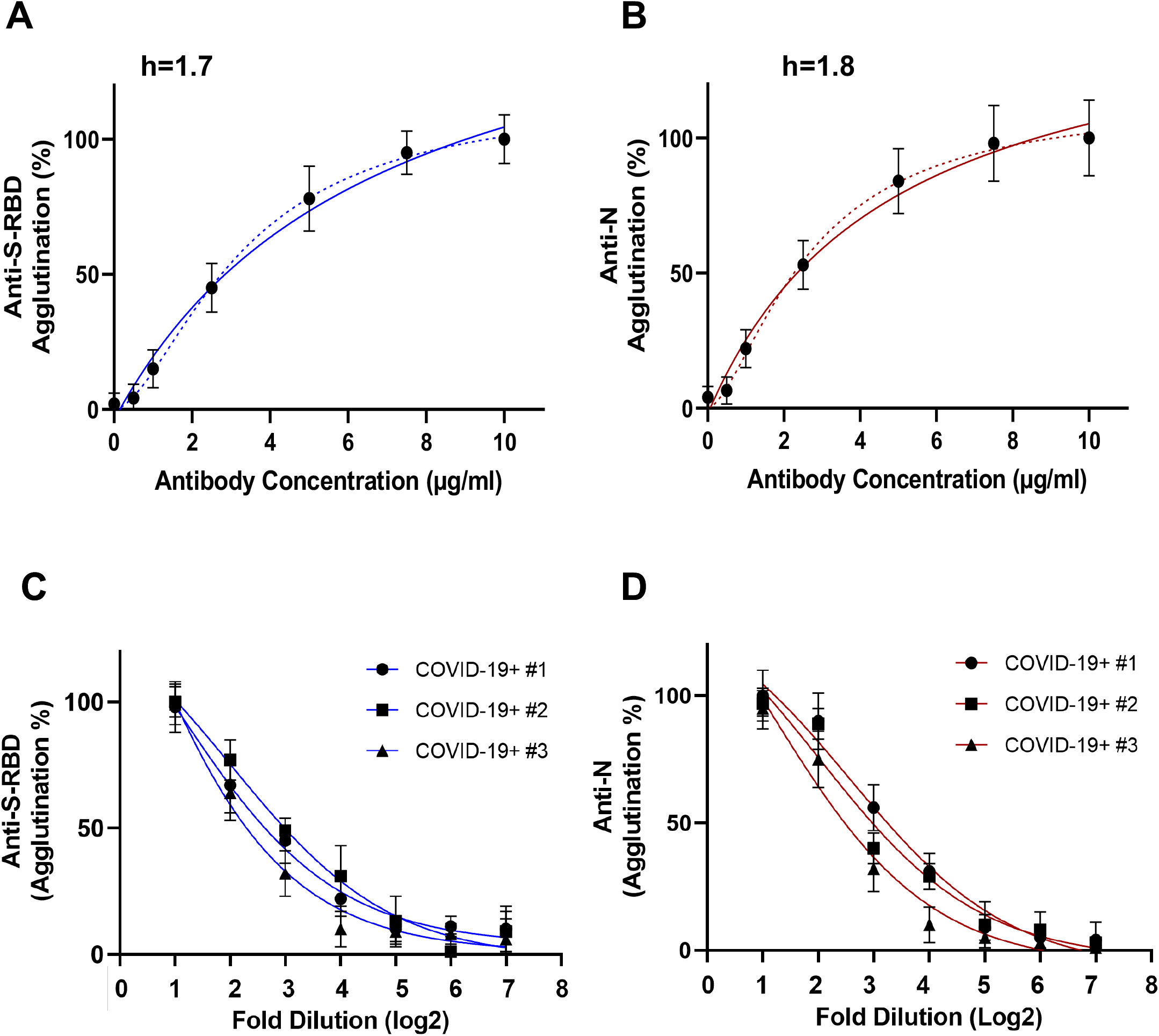
Antibody-induced latex particle agglutination correlates with the antibody titer. (**A, B**) Changes in agglutination in response to increased concentrations of the anti-S-RBD (n=3) or anti-N antibody (n=3). Dash lines represent fitted curves to Hill equation (h, Hill coefficient). (**C, D**) S-RBD (C) or N (D) antibody-induced agglutination decreased with increased dilution of plasma. Shown are agglutination data (in log2 scale) from three COVID-19^+^ plasma samples with 1:2 to 1:128 dilution. Data shown are from three replicates per concentration.

We next examined if the latex agglutination assay could be used to gauge COVID-19 antibody response. Using plasma samples from patients tested positive for SARS-CoV-2 by NAAT and confirmed for strong antibody response by ELISA, we found that the patient plasma samples were not only capable of inducing agglutination of the S-RBD- or N-coated latex particles, they did so in a concentration-dependent manner. As shown in Fig. 2C&D, the extent of agglutination decreased as the plasma was diluted, indicating that the agglutination assay may be used to estimate antibody titer as in an ELISA-based antibody test.

### The latex agglutination-based antibody assay showed high sensitivity and specificity

To validate the antibody test based on latex particle agglutination, we carried out agglutination assays for 290 residual plasma samples from individuals that were tested positive (169) or negative (121) for virus RNA by the Roche cobas SARS-CoV-2 test^9^. To assess specificity, we also included 100 virus-naïve samples banked in 2018 in our agglutination assay. None of the 121 SARS-CoV-2^-^ or the 100 Pre-COVID-19 plasma samples was capable of promoting the agglutination of either the S-RBD- or N-coated latex particles, indicating 100% specificity for the agglutination assay (Table 1). In contrast, of the 169 SARS-CoV-2^+^ plasma samples tested, 166 (98.2%) promoted agglutination in response to the S-RBD antigen and 164 (97%) to the N antigen, with overall sensitivity of 98.2%. We compared the latex agglutination assay with the Euroimmune IgG test for the S antibody and the Roche Elecsys® Total assay for the N antibody using the same set of SARS-CoV-2^+^ plasma samples and found that the latex agglutination assay outperformed both ELISA-based antibody tests (Table 2). The agglutination assay also exhibited better specificity than either ELISA kit (Table 2). Quantification of the agglutination data showed that the COVID-19^+^ group is significantly different from the COVID-19^-^ or pre-COVID-19 group, indicating that the latex agglutination assay effectively distinguished SARS-CoV2^+^ from SARS-CoV2^-^ individuals (Fig. 3A&B).

**Table 1:** Clinical performance of the agglutination-based antibody assay.

| Samples | Anti-S-RBD<br>(n) |  | Anti-N<br>(n) |  | Overall<br>Sensitivity | Overall<br>Specificity |
| --- | --- | --- | --- | --- | --- | --- |
|  | Positive | Negative | Positive | Negative |  |  |
| SARS-CoV-2 NAAT<br>positive (n=169) | 166 | 3 | 164 | 5 | **98.2%**<br>(166/169) |  |
| **Days of SARS-CoV-2**<br>**NAAT positive** |  |  |  |  |  |  |
| 1<br>(n=41) | 38 | 3 | 36 | 5 | 92.7%<br>(38/41) |  |
| 2<br>(n=31) | 30 | 1 | 31 | 0 | 100% |  |
| ≥3<br>(n=97) | 97 | 0 | 96 | 1 | 100% |  |
| **SARS-CoV-2 NAAT**<br>**negative**<br>**(n=121)** | 0 | 121 | 0 | 121 |  | **100%** |
| **Pre-COVID-19**<br>**(n=100)** | 0 | 100 | 0 | 100 |  | 100% |

**Table 2:** Comparison in sensitivity and specificity between the agglutination-based and ELISA-based antibody assays.

| Anti-S (RBD) | Agglutination assay<br>(n) |  | ELISA<sup>9</sup><br>EUROIMMUNE<br>(n) |  | Sensitivity<br>(Agglutination) | Sensitivity<br>(Euroimmune) | Specificity<br>(Agglutination) | Specificity<br>(Euroimmune) |
| --- | --- | --- | --- | --- | --- | --- | --- | --- |
|  | Positive | Negative | Positive | Negative |  |  |  |  |
| SARS-CoV-2 NAAT<br>positive (n=121) | 118 | 3 | 90 | 31 | 97.5%<br>(118/121) | 74.4%<br>(90/121) | NA |  |
| SARS-CoV-2 NAAT<br>Negative &<br>preCOVID-19 | 0 | 221 | 11 | 194 | NA |  | 100%<br>(221/221) | 94.6%<br>(194/205) |
| Anti-N | Agglutination assay<br>(n) |  | ELISA<sup>9</sup><br>Roche Elecsys®<br>(n) |  | Sensitivity<br>(Agglutination) | Sensitivity<br>(Roche) | Specificity<br>(Agglutination) | Specificity<br>(Roche) |
|  | Positive | Negative | Positive | Negative |  |  |  |  |
| SARS-CoV-2 NAAT<br>positive (n=69) | 68 | 1 | 66 | 2 | 98.6%<br>(68/69) | 95.7%<br>(66/69) | NA |  |
| SARS-CoV-2 NAAT<br>Negative &<br>preCOVID-19 | 0 | 221 | 1 | 135 | NA |  | 100%<br>(221/221) | 99%<br>(135/136) |

**Figure 3.**
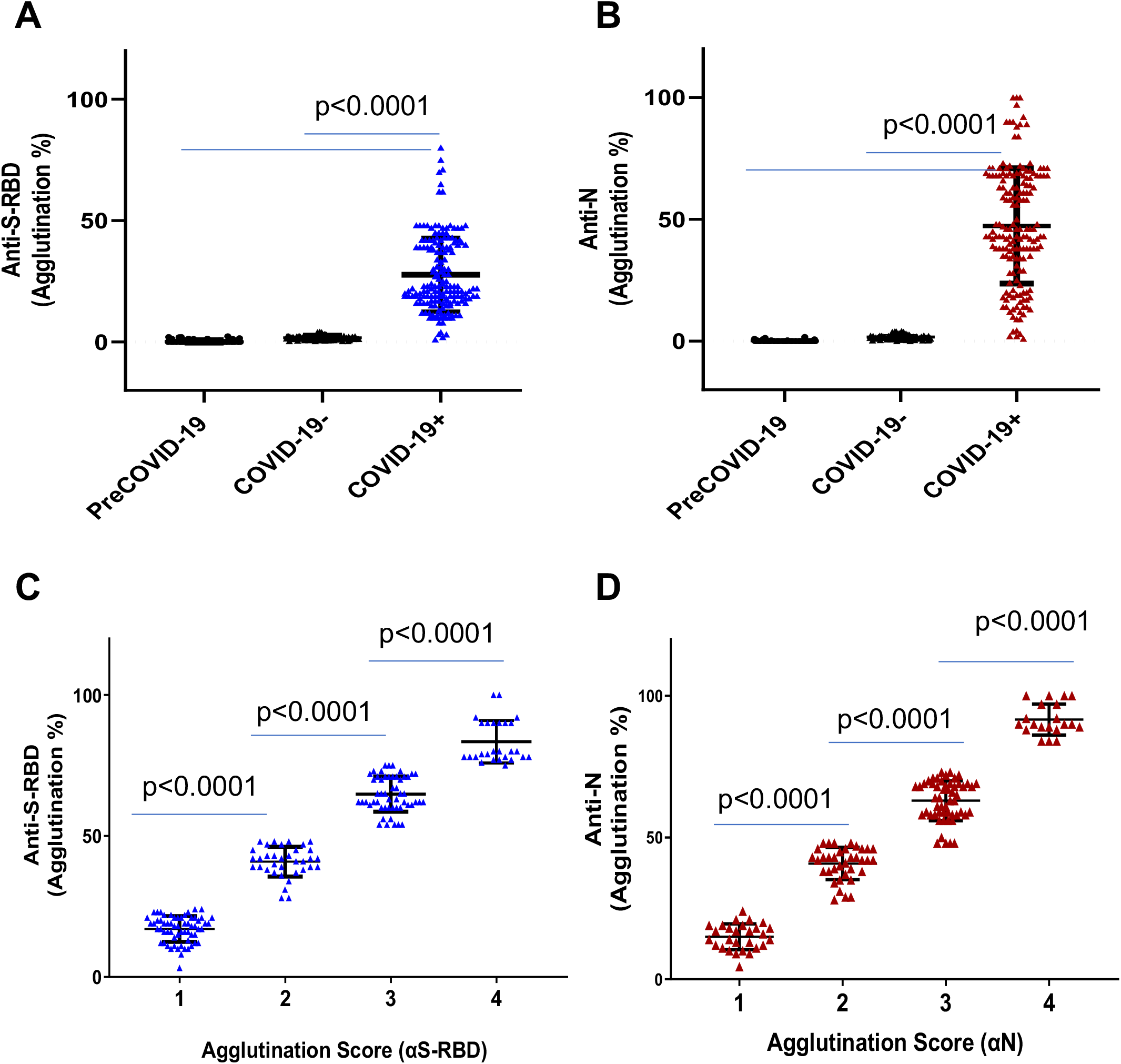
Agglutination assay distinguished COVID-19^+^ from COVID-19^-^ samples. (**A, B**) Comparison of S-RBD (A) and N antibody responses between COVID-19^+^ (n=169), COVID-19^-^ (n=121) and Pre-COVID-19 (n=100) samples determined by the aggregation assay. (**C, D**) The strength of the S-RBD and N antibody response in the COVID-19^+^ (n=169) plasma samples could be determined semi-quantitatively by the aggregation score (1-4 denotes weak-to-strong antibody response). Statistical analyses were performed using unpaired Student’s t-test with Welch’s correction (p values shown on graph).

Based on the background signals of the COVID-19^-^ samples (0-4% agglutination for both S-RBD- and N-coated latex particles), we set 5% agglutination as the cut-off for antibody positivity. To facilitate the use of the latex agglutination assay as a simple, semi-quantitative antibody test, we developed a numerical scoring system for antibody response. We assigned the scores 1, 2, 3 and 4 to samples that produced 5-25%, 25-50%, 50-75% and >75% agglutination, respectively (Fig. S4). We found that this scoring scheme effectively distinguished samples with strong antibody response from those with medium or weak ones (Fig. 3C&D). The agglutination score may be readily assigned by visual inspection and comparing to reference wells containing a predetermined amount of pure anti-S-RBD or anti-N antibody (Fig. S3).

### The S-RBD antibody response correlated with neutralizing antibody titer

Because neutralizing antibodies (Nab) play a pivotal role in the humoral immune response to the virus^17^, we next examined if the S-RBD antibody response determined by the agglutination assay correlated with neutralization efficiency. We developed a surrogate neutralization assay by measuring the efficacy of patient plasma in blocking S-RBD binding to its host receptor, angiotensin converting enzyme 2 (ACE2) *in vitro*. Similar approaches have been used by others to evaluate neutralization efficiency of patient plasma or therapeutic antibodies^18,19^. Briefly, binding of biotinylated ACE2 to immobilized S-RBD is detected by ELISA through HRP-conjugated streptavidin. The presence of neutralizing antibody would block this interaction, resulting in reduction of the ELISA signal. Using this surrogate neutralization assay, we found that the neutralization efficiency increased with the agglutination score for the S-RBD antibody (Fig. 4A). Intriguingly, comparison of samples with distinct S-RBD and N antibody responses indicated that the neutralization efficiency was significantly correlated with the S-RBD, but not the N antibody strength. This is not surprising given that the nucleocapsid is not involved in mediating virus entry into the host cells via ACE2. Nevertheless, it remains to be determined if the plasma with strong N antibody response would confer immunity by inhibiting virus replication *in vivo*.

**Figure 4.**
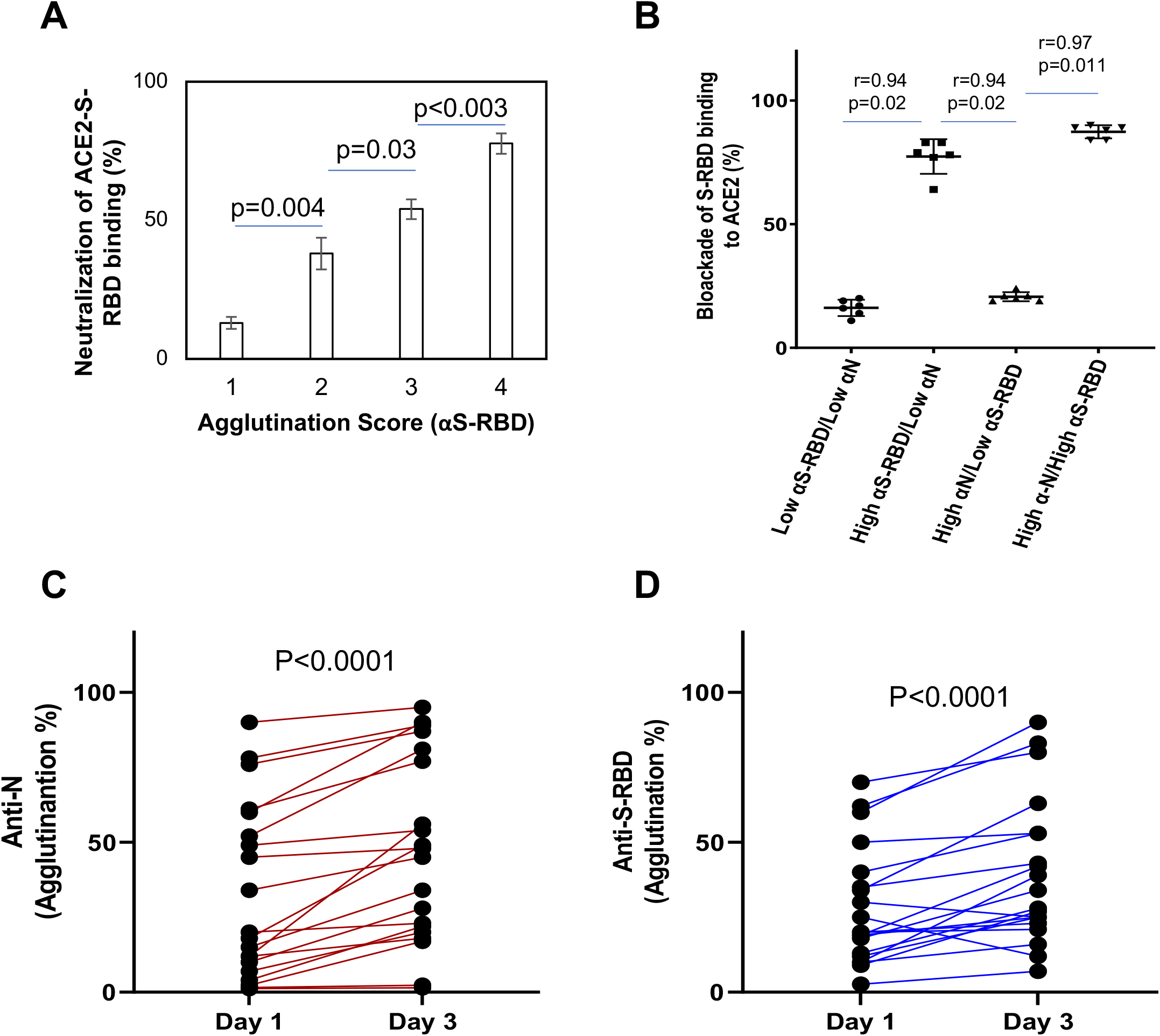
Using the latex agglutination assay to determine neutralizing antibody titer and dynamic changes in antibody response. **(A)** Neutralization antibody response correlated significantly with the agglutination score. P values calculated based on unpaired Student t-test (n=10 for each group). (**B**) Spearman (r) correlation of efficiency of neutralization and S-RBD or N antibody response (n=25). (**C, D**) Dynamic changes in antibody responses in COVID-19 patients. n=20 for day 1 and n=20 for day 3; p-values calculated based on unpaired Student’s t-test with Welch’s correction.

### The agglutination assay allowed for early antibody detection and tracking of dynamic antibody response

We noted that the agglutination assay detected antibody response in >92% plasma samples collected on the day of SARS-CoV2^+^ diagnosis by NAAT and in 100% samples on day 2 and afterward. This is in stark contrast to the 47% to 83% sensitivity for ELISA-based antibody tests on samples collected within 7 days of positive NAAT^9^. The superb sensitivity of the latex agglutination assay suggests that it may be used to detect antibody response in the early stage of virus infection and monitor its dynamic changes over time^12,20,21^. Using serial blood samples from SARS-CoV-2^+^ patients, we compared the changes in the S-RBD and N antibody responses between days 1 and day 3 (Fig. 4C&D). We found that the majority of patients showed detectable S-RBD and/or N antibodies on NAAT^+^ day 1. Moreover, the antibody titer increased significantly on day 3 compared to day 1. Because it is not possible to determine how long these patients had contracted the virus prior to the NAAT test, seroconversion might have occurred for some on the day of diagnosis. Nevertheless, we were able to detect anti-S-RBD or anti-N antibodies on day 3 for several patients who showed no detectable antibody on day 1, suggesting seroconversion occurred rapidly in these patients^4^. Taken together, these data suggest that the latex agglutination assay may be used to diagnose active infection in conjunction with NAAT. Combined antibody and RNA testing may increase the sensitivity of the later. Moreover, the agglutination-based antibody test may be used to monitor the evolvement of humoral immune reaction in infected individuals over time.

## DISCUSSION

Antibody testing offers an additional and much needed tool for managing the COVID-19 pandemic, which may allow for rapid and cost-effective POC diagnosis to facilitate treatment and public health responses. Furthermore, antibody testing may play an important role in identifying individuals who have gained protective immunity from previous exposure or immunization programs. While the clinical trial results for several vaccine candidates are encouraging, seroconversion is unlikely to occur for all vaccinated individuals^22^. A highly sensitive and specific antibody test would allow for accurate assessment of humoral response to a vaccine, and serial testing post-vaccination could indicate the duration of humoral immunity against SARS-CoV-2. Such information will be invaluable to inform public health decisions, both for directing resource allocation and determining response to vaccination.

While most serology tests used in the clinic are specific for a specific antibody isoform, IgG, IgM or IgA, the latex agglutination antibody assay is, in principle, isotope-independent^11^. This may contribute to its high sensitivity, especially on samples collected in the early phase of infection when IgG isotope switching has not yet occurred. It has been shown that the IgM antibody response proceeds that of IgG^3,11^, and the pentameric architecture of IgM may make it a stronger promoter of agglutination than IgG. Moreover, the agglutination assay detects antibodies against the spike (S) and the nucleocapsid (N) in parallel whereas most existing assays are specific for either protein. These factors explain why the agglutination assay could detect antibody in >92% plasma samples collected on the day of NAAT^+^ diagnosis and 100% on day 2 and onwards whereas other antibody tests were less sensitive. The ability of our assay to detect antibodies at the early stages of infection suggests that it may be used to complement or confirm the diagnosis based on NAAT, which is prone to false positives or false negatives especially when done only once^23^.

Several features of the agglutination assay make it an ideal candidate for POC or at-home antibody testing. First, the assay is highly sensitive and accurate, with 100% specificity and ∼98% sensitivity on the samples tested in this study. Importantly, it detected antibodies in >92% COVID-19 patients on the day of diagnosis. This sensitivity rivals that of NAAT tests^23^. Second, the agglutination test is fast. It takes two minutes from mixing the plasma with the latex particles to getting the result. Third, the agglutination assay is simple to run and instrument-free. As the formation of latex bead clumps is easy to identify by the naked eye, the test may be performed without extensive specialized training. Fourth, because the agglutination assay takes only ∼5 µl plasma, it may be developed into a finger prick blood test suitable for at-home use. Finally, the low cost of the latex agglutination assay makes it universally affordable and ideal for antibody testing at the community or population level, in high- and low-resource settings alike.

## Data Availability

All data will be made available upon request

## ACKNOWLEDGEMENT

We thank the Center for Structural Genomics of Infectious Diseases (CDGID) for the N-RBD expression plasmid, Dr. Jason McLellan for expression plasmid for human ACE2, Dr. Shane Harding for the spike-receptor binding domain expression construct, and Yanjun Li for technical assistance. This work was supported by a grant (to SSCL, ICY, MK, BCY and VB) from the Ontario Research Fund-COVID-19 Rapid Research Fund and by the Toronto COVID-19 Action Fund. SE was supported by a Post-Doctoral Fellowship from the National Science and Engineering Council of Canada. SSCL held a Canada Research Chair in Molecular and Epigenetic Basis of Cancer.

## MATERIALS AND METHODS

### Blood sample collection

Blood samples were collected following a protocol (study number: 116284) approved by the Research Ethics Board (REB) of Western University. The plasma samples were de-identified prior to transfer from the Laboratory of Clinical Medicine (London Health Sciences Center, London, Canada) to a biosafety Level 3 (CL3) lab (ImPaKT, Western University) following Transportation of Dangerous Goods (TDG) guidelines. All plasma samples were heat-inactivated at 56 ^°^C for 30 minutes at the ImPaKT CL3 facility as per Western university biosafety regulation. Heat inactivated plasma samples were then transferred to the testing laboratory. We tested the effect of heat inactivation on SARS-CoV-2 antibody titer and found no significant impact of heat-inactivation (see also Fig. S5).

### Recombinant protein production and purification

The expression plasmid for human ACE2 cloned into the mammalian expression vector pαH (residues 1−615 with a C-terminal HRV3C protease cleavage site, a TwinStrepTag and an 8XHisTag) was a generous gift by Dr. McLellan. SARS-CoV-2 S-RBD cloned into pCAGGs the expression vector was received from Dr. Harding’s lab. SARS-CoV-2 N-RBD was cloned into the pMCSG53 prokaryotic expression vector (residues 47-173-terminal 6x-His tag + TEV protease cleavage site).

Recombinant ACE2 and S-RBD proteins were produced by transient transfection of Expi293F cells (ThermoFisher Scientific, A14527) with a corresponding expression vectors and FectoPRO® DNA transfection reagent (Polyplus-transfection® SA, Cat. #116-010). Supernatants from transfected cells were harvested after 96 hours of the post-transfection time by centrifugation of the culture at 65,000 RPM for 30 min at 4^°^C. Cleared supernatant was then incubated with 4 ml TALON® Metal Affinity Resin (Takara Bio USA, Inc. Cat#635652) for 2h at 4^°^C. Ni-NTA chromatography was used to purify His-tagged recombinant proteins. Each protein was dialyzed and concentrated in Amicon centrifugal units (EMD Millipore) in a final buffer of 20mM HEPES (pH 7.5, 200mM NaCl, pH 7.5, 5% glycerol).

Recombinant N-RBD was expressed in *E. coli* BL21(DE3)-Gold. Ni-NTA chromatography and size exclusion chromatography-Superdex S200 was used to purify N-RBD. The tag was cleaved using TEV followed by dialysis. The N-RBD protein was resuspended in 0.1 M NaCl, 20 mM HEPES pH 7.5 and stored at −80°C until use. Recombinant nucleocapsid (residues 1-419) was obtained from RayBiotech (Cat #230-01104). Protein purity was confirmed by SDS-PAGE (Fig. S1).

### Preparation of SARS-CoV-2 antigen coated latex particles

Blue dyed polystyrene latex beads, 0.8 *µ*m in diameter, were purchased from Sigma Aldrich (L1398). Prior to use, the latex beads were washed according to the manufacturer’s instructions with some modifications. Briefly, 2.5mL of 5% (w/v) latex suspension was washed twice in 10 mL PBS buffer (135 mM NaCl, 2.6 mM KCl, 8 mM Na_2_HPO_4_, and 1.5 mM KH_2_PO_4_, pH 7.4) by mixing and centrifuging the latex suspension at 3,000g for 10 minutes at room temperature. The beads were then resuspended with 2.5 ml 0.025M MES buffer (2-(N-Morpholino) ethanesulfonic acid, pH 6.0) to obtain 5% (w/v) suspension.

SARS-CoV-2 antigen-latex particle conjugates were prepared by passive adsorption following the procedures described by Mahat et al^24^, with some modifications. Briefly, 0.4 mL of 5% (w/v) latex suspension was centrifuged at 3,000 g for 5 minutes at room temperature, and the supernatant was discarded. The beads were incubated with 200 µg recombinant Receptor Binding Domain of the SARS-CoV-2 spike protein (S-RBD) (Structural Genomics Consortium, University of Toronto) or the Nucleocapsid protein (N protein) (RayBiotech, 230-01104) in 4 mL MES buffer. The mixture was allowed to incubate for 24 hours at 4°C with periodic mixing. After conjugation, the antigen-latex bead conjugate was centrifuged, and the supernatant was kept for determination of unabsorbed protein concentration (Bio-Rad protein assay kit). The antigen-bead conjugate was washed twice with PBS and blocked for 30 min at room temperature in PBS containing 3% bovine serum albumin (BSA). The conjugate was then resuspended at 2.5% (w/v) in PBS containing 1% BSA and stored at 4°C until use.

### Agglutination assay for SARS-CoV-2 antibody testing and data interpretation

For the agglutination assay, 5 µl plasma was mixed with 25 µl antigen-coated beads (2.5%, w/v) per assay. The agglutination was allowed to proceed for 2 min at room temperature before imaging with a camera. The relative degree of agglutination induced by the SARS-CoV-2 antibody was measured by the area of clump formation based on the corresponding image. Agglutination data analyses were performed using qualitative and semi-quantitative assessments. For semi-quantification of agglutination, the image analysis software Qupath (v0.1.2) was used (https://qupath.github.io/) and quantification was done by calculating the percentage of agglutination based on estimated agglutination/clumps area (mm^2^) relative to the total latex reaction area. In qualitative assessments, agglutination intensity was inspected visually, and agglutination score was assigned (i.e. 1, 2, 3 and 4). Specifically, 1 corresponds to small clumps with ∼25% agglutination, 2 (∼50% agglutination), 3 (∼75% agglutination), and 4 (large clumps that forms in less than 1 min with ∼100% agglutination). Based on data from the COVID-19 negative samples (including NAAT negative and samples collected in 2018), the cut-off for positivity was set to 5% of agglutination.

### Preparation of red blood cells conjugated with SARS-CoV-2 antigen

The recombinant spike receptor-binding domain (S-RBD) or the nucleocapsid RNA-binding domain (N-RBD) was conjugated in 30-fold molar excess biotin using EZ-Link Sulfo-NHS-LC-LC-Biotin (Thermo Scientific, A35358). Excess unbound biotin was removed using ZebaTM Spin Desalting Columns, 7KMWCO (Thermo Scientific, 89890). Anti-D-IgG was purified from Immucor Anti-D Series 4 (IgG & IgM monoclonal blend) by using protein A magnetic affinity purification (G8782, Promega). The purified anti-D-IgG was then concentrated (3mg/ml) and stored at 4 °C until use. Anti-D was then conjugated with streptavidin according to manufacturer instruction (ab102921, abcam).

Bioconjugation of Anti-D-IgG-streptavidin with Reagent Red Blood Cells (RRBC) [0.8% R2R2; blood group O; Rh/D-antigen+] (Ortho-Clinical Diagnostics SELECTOGEN, 6902315) was done by incubating the anti-D-IgG-streptavidin with RRBC for 30 min at room temperature. The RRBC-anti-D-streptavidin complex was then washed twice with low ionic strength RBC diluent (MTS™ Diluent 2 PLUS; Micro Typing Inc., MTS9330S). The complex was centrifuged at 1000g for 2 min to remove unbound anti-D-IgG streptavidin and was then resuspended in the same RBC diluent. RBC-anti-D-IgG-streptavidin was then conjugated with either biotin-S-RBD or biotin-N-RBD for 15 min at room temperature. The RRBC-anti-D-sterptaviding-biotin-S-RBD/N-RBD was stored at 4 °C until use. The RRBC agglutination assay was carried out in the same way as for latex agglutination described above.

### S-RBD-ACE2 binding ELISA and surrogate neutralization assay

#### ELISA plate Coating and blocking

S-RBD was dissolved (5 µg/ml) in Tris buffer saline (TBS) (20 mM Tris, 150 mM NaCl, pH7.4) and 100 µl of the S-RBD solution was added to each well of an ELISA plate and incubate at 4°C overnight with slow shaking. The antigen-coated wells were washed 3 times with TBS-tween (TBST) (20 mM Tris, 150 mM NaCl, 0.1% Tween 20). The S-RBD coated wells were blocked by 100 µl of the ChonBlock^™^ blocking/sample dilution ELISA buffer (Chondrex, Inc., 9068) for 1 hour at room temperature with slow shaking followed washing 3 times with TBST.

#### ACE2:S-RBD binding assay

ACE2 was biotinylated as described above. Biotin-ACE2 (1µg/m) was added to S-RBD-coated plate after blocking and incubated for 1hour at room temperature. The wells were washed 3 times with TBST to remove unbound biotin-ACE2. Streptavidin-HRP (1000-fold dilution with Chonblock blocking buffer) was then added to each well and incubated for 1hour at room temperature. The wells were washed 3 times with TBST and TMB substrate (3,3’,5,5’-Tetramethylbenzidine, Thermo Scientific, N301) was added for reaction development and 0.18 M H2SO4 was used to stop reaction. Absorbance at 450nm was measured to detect the S-RBD bound ACE2.

#### SARS-CoV-2 antibody neutralization assay

Plasma was diluted 1:100 and incubated with S-RBD-coated wells (blocked) for 1hour at room temperature. The wells were washed three times with TBST. Biotin-ACE2 was then added to the wells and incubated for 1 hour at room temperature followed by washing, reaction development and detection as described above.

### Statistical analysis

All statistical analyses were done using the GraphPad Prism9 software. Specifically, the Hill coefficient (h) was calculated from fitting agglutination data obtained using anti-S-RBD and anti-N antibodies to the Hill equation. COVID-19^+^ samples with distinct agglutination scores and COVID-19^-^ samples were analyzed using unpaired t-test with Welch’s correction (no assumption of equal SD between two groups). Changes in agglutination for samples before and after heat-inactivation were analyzed by paired t-test. Spearman’s correlation rank was done to study correlation between antibody titter and ACE2:S-RBD neutralization efficiency.

## Supplementary Figures

**Figure S1.**
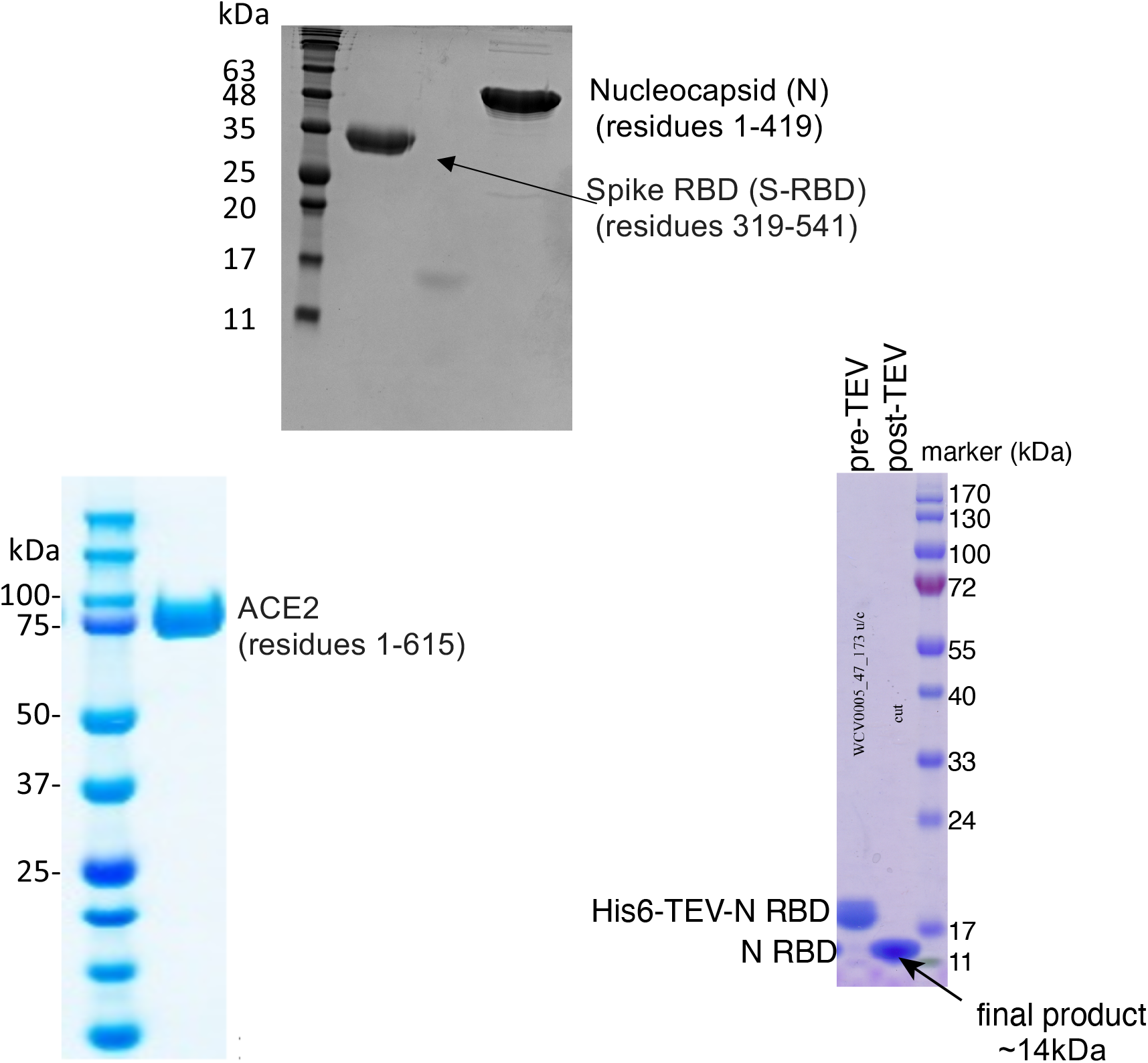
SDS-PAGE images of recombinant SARS-CoV-2 proteins employed in the current study.

**Figure S2.**
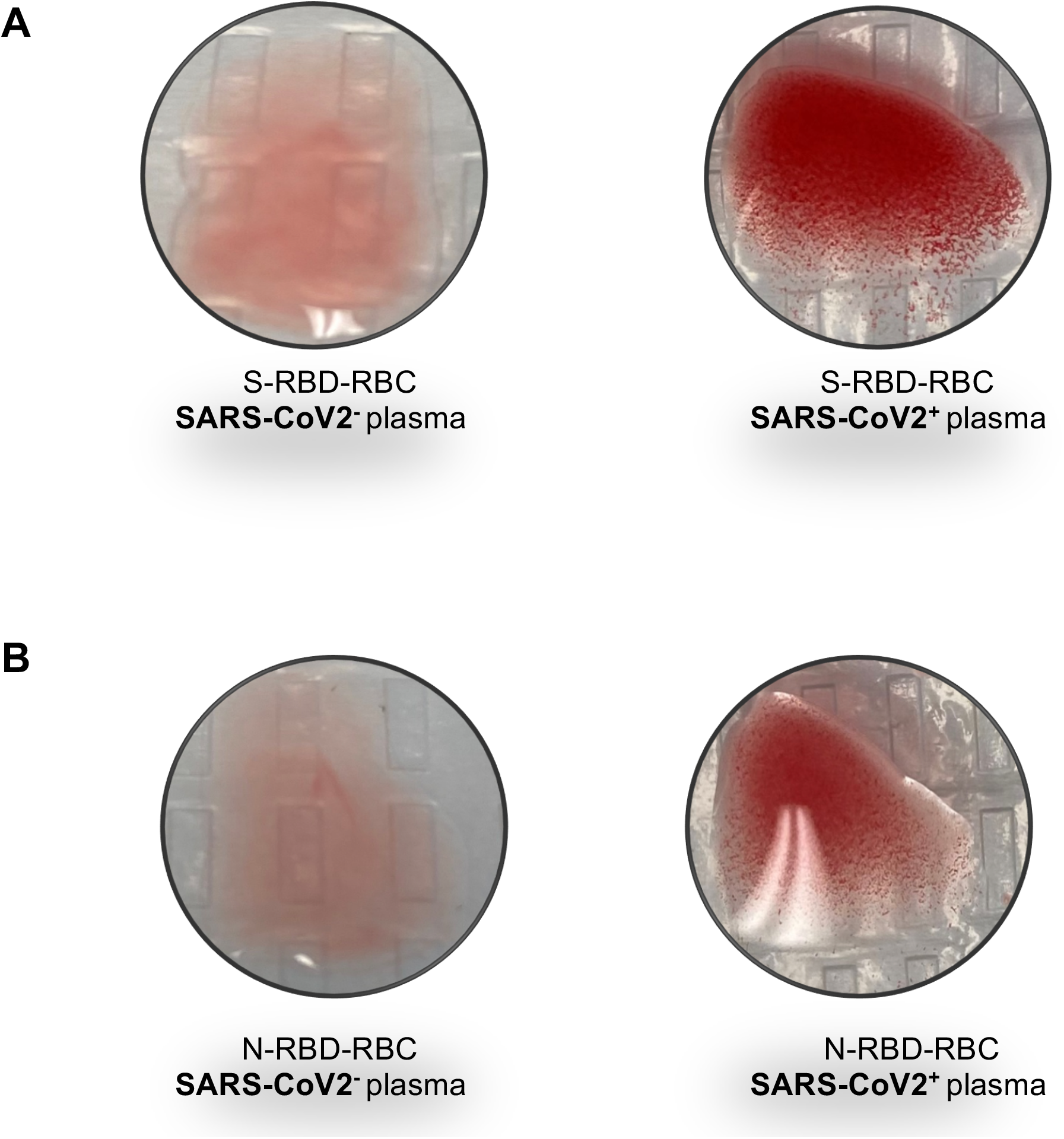
SARS-CoV2 antibody testing based on RBC agglutination. Red blood cells (RBC, group O; R2R2) carrying the D antigen were labelled with anti-D IgG conjugated to recombinant S-RBD or N-RBD through streptavidin-biotin (i.e., IgG-streptavidin conjugated to biotin-RBD). (A) S-RBD labeled RBCs were mixed with either SARS-CoV-2^-^ (NAAT) or SARS-CoV2^+^ plasma (right). (B) N-RBD labeled RBCs were mixed with either SARS-CoV2^-^ (left) or SARS-CoV2^+^ plasma (right). Images shown were taken after 1 min incubation at room temperature.

**Figure S3.**
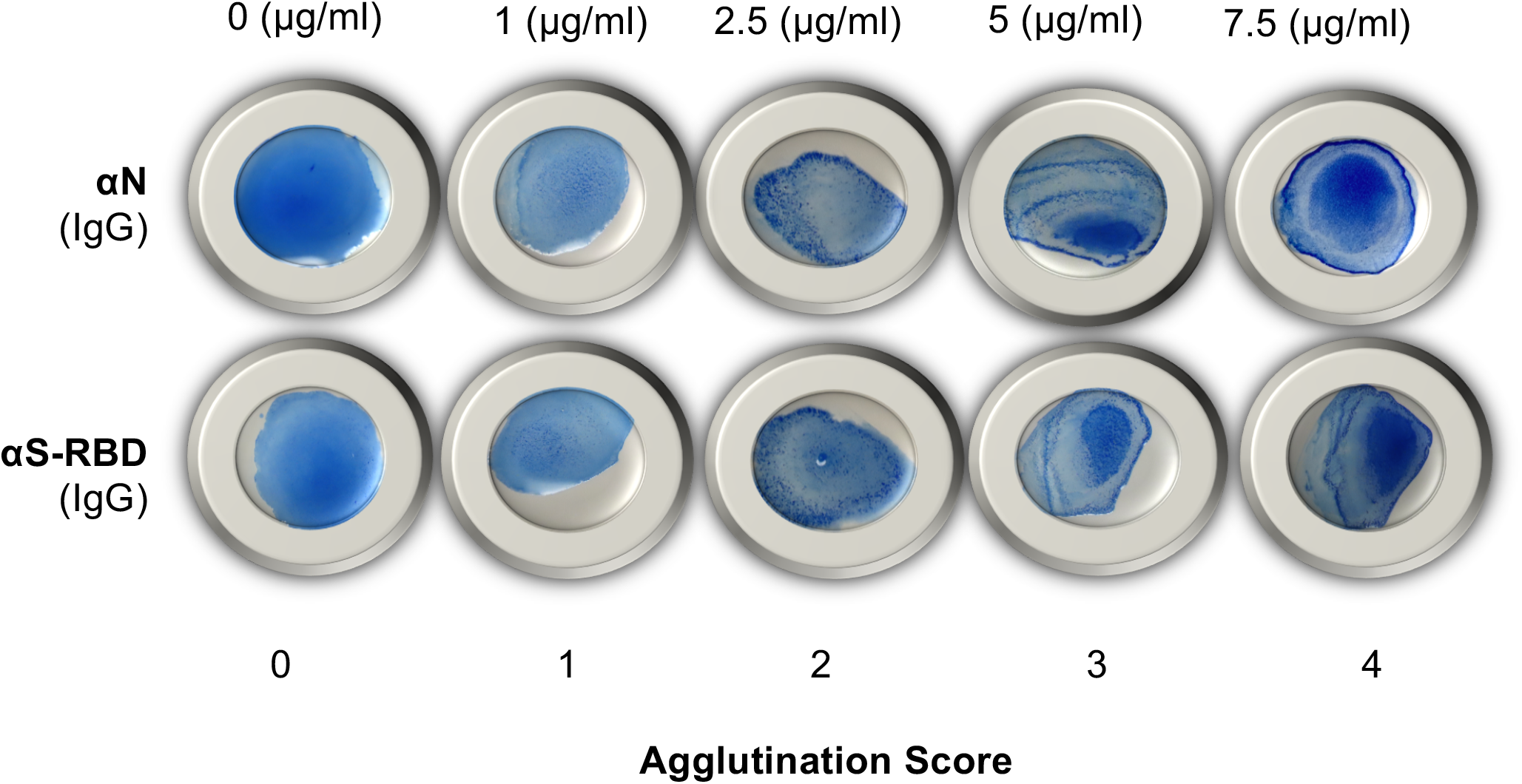
Agglutination of blue latex particles in response to different concentrations of antibody (IgG) against the nucleocapsid (*α*N) or S-RBD (*α*S-RBD). Anti-S-RBD (monoclonal, NBP2-90980) was obtained from Novus Biologicals; Anti-Nucleocapsid (polyclonal, PA5-81794) was from ThermoFisher Scientific.

**Figure S4.**
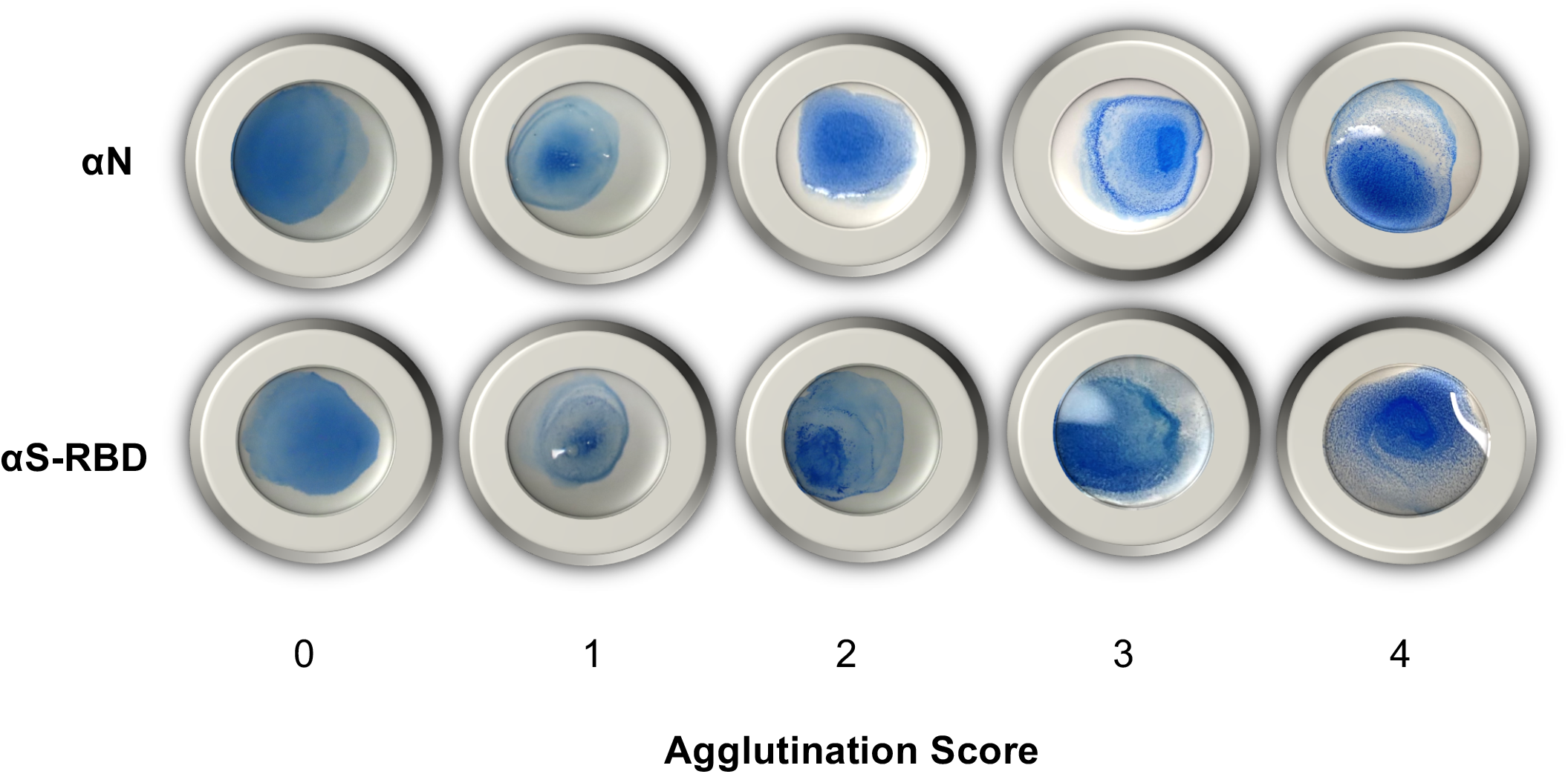
Representative images of agglutination induced by plasma with different agglutination scores. Shown are five samples with distinct agglutination scores (0-4) in the nucleocapsid (*α*N) (upper row) or the S-RBD antibody (*α*S-RBD) test (lower row). The scores were assigned as 4 = 75-100% agglutination; 3 = 50-75% agglutination; 2 = 25-50% agglutination; 1 = 5-25% agglutination; 0 (or negative) ≤5% agglutination.

**Figure S5.**
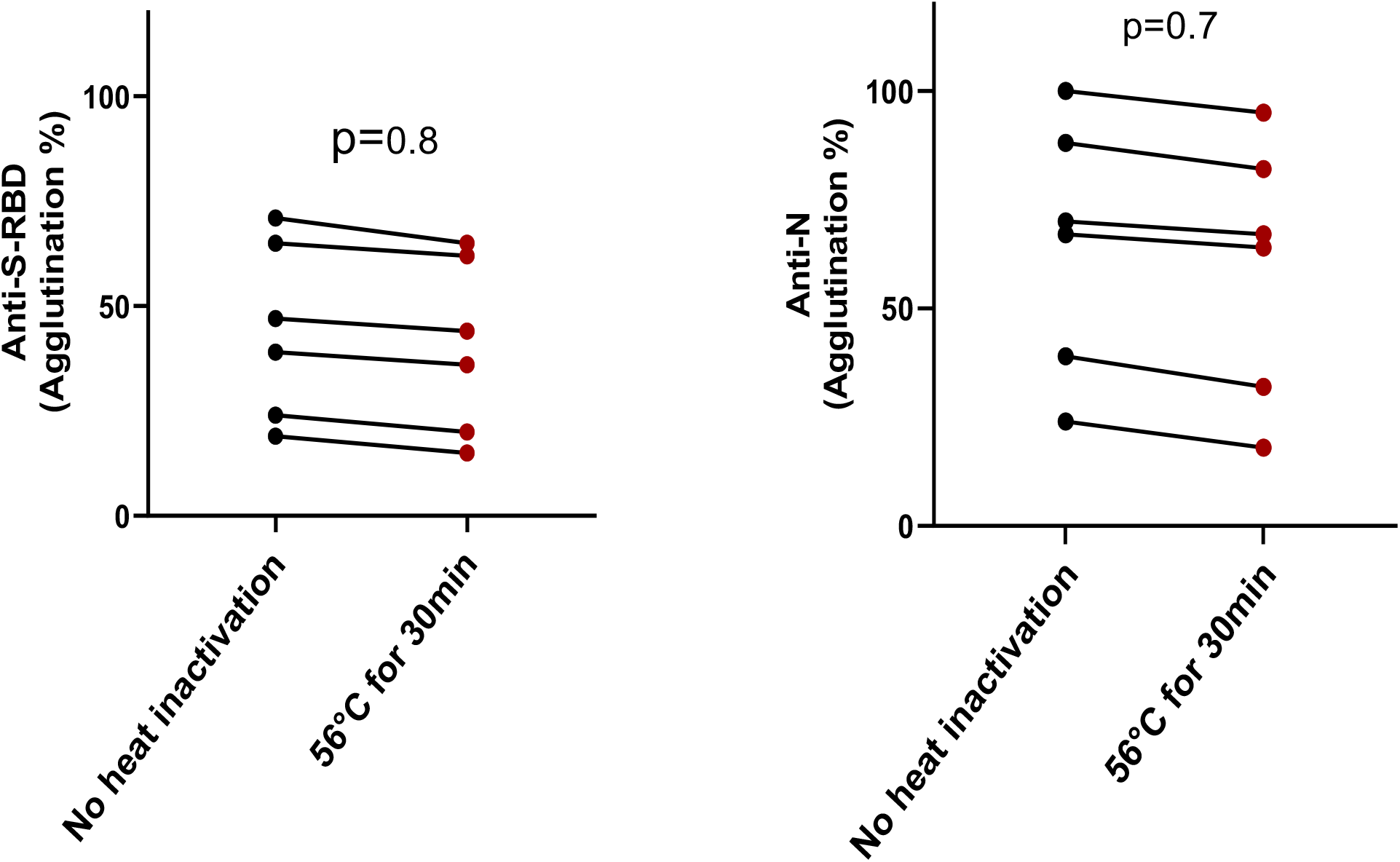
Heat inactivation of plasma did not affect antibody induced agglutination. Shown are agglutination percentages between samples without or with heat-inactivation in the anti-S-RBD (left) or anti-N (right) agglutination assay. p-values calculated from paired two tailed t-test (no assumption of equal variance, n=6).

## Notes

### Competing Interest Statement

SE, CV and SSCL are inventors in a provisional patent submitted by Western University

### Author Declarations

Blood samples were collected following a protocol (study number: 116284) approved by the Research Ethics Board (REB) of Western University. The plasma samples were de-identified prior to transfer from the Laboratory of Clinical Medicine (London Health Sciences Center, London, Canada) to a biosafety Level 3 (CL3) lab (ImPaKT, Western University) following Transportation of Dangerous Goods (TDG) guidelines. All plasma samples were heat-inactivated at 56 oC for 30 minutes at the ImPaKT CL3 facility as per Western university biosafety regulation. Heat inactivated plasma samples were then transferred to the testing laboratory.

